# Which countries need COVID-19 vaccines the most? Development of a prioritisation tool

**DOI:** 10.1101/2022.04.27.22274377

**Authors:** Vageesh Jain, Rifat Atun, Paul Hansen, Paula Lorgelly

## Abstract

**Background:** Assessing relative needs for COVID-19 vaccines across countries has been challenging. The objective of this study was to identify the most important factors for assessing countries’ needs for vaccines, and to weight each, generating a scoring tool for prioritising countries.

**Methods:** The study was conducted between March and November 2021. The first stage involved a Delphi survey with a purposive and snowball sample of public health experts, to reach consensus on country-level factors for assessing relative needs for COVID-19 vaccines. The second stage involved a discrete choice experiment (DCE) to determine weights for the factors.

**Results:** The study included 28 experts working across 13 different countries and globally. The Delphi survey found 37 factors related to needs. Nine of the most important factors were included in the DCE. Among these, the most important factor was the ‘proportion of overall population not fully vaccinated’ with a mean weight of 19.5, followed by ‘proportion of high-risk population not fully vaccinated’ (16.1), ‘health system capacity’ (14.2), ‘capacity to purchase vaccines’ (11.9) and the ‘proportion of the population clinically vulnerable’ (11.3).

**Conclusions:** By assessing relative needs, this scoring tool can build on existing methods to further the role of equity in global COVID-19 vaccine allocation.

## Introduction

The equitable global allocation of COVID-19 vaccines has received considerable attention, although to date the concept of an ‘equitable allocation’ of vaccines has been poorly defined. Understanding vaccine equity requires an assessment of the need for vaccines across countries. This subject pertains to vertical equity – how resources are prioritised among those with varying needs – as well as horizontal equity, where countries with similar needs for vaccines should have comparable levels of access.

The COVID-19 pandemic and associated non-pharmaceutical interventions (NPIs) have affected countries and populations differently, making needs assessment complex. Nevertheless, the global allocation of vaccines has been most closely aligned with countries’ ability-to-pay and vaccine manufacturing capacity rather than needs (1). Despite the rapidity of clinical trials and the accelerated production of vaccines, six months after the first approval of a Covid-19 vaccine, only 1% of people in low-income countries had been vaccinated, compared to 43% in high-income countries (1).

Current methods for allocating vaccines between countries are narrow in scope (2-5). The COVAX Facility aims to allocate vaccines to cover 20% of each national population, followed by a needs assessment that considers a small range of metrics and a qualitative assessment (6). Although established with the ambition to support the equitable distribution of resources in a global emergency, the COVAX Facility has suffered from insufficient access to vaccines. The arrival of the Omicron variant led to high-income countries initially administering more booster doses than all vaccine doses combined in low-income countries (7). Under such resource constraints, the process used to allocate scarce vaccines across countries, which was hastily designed during a global health crisis, warrants scrutiny and, if possible, improvement.

We recently proposed a conceptual framework (COVID-NEEDS) (8) that considered vaccine needs to be affected by a wide range of health, social and economic impacts of COVID-19 and associated NPIs. The framework’s usefulness has so far been hampered by an inability to validate the proposed factors or consider the relative importance of the factors included.

The objective of the present study was to identify the most important factors for assessing countries’ relative needs for COVID-19 vaccines, and to establish weights for them to create a scoring tool. With persistent inequities and the potential need for further doses in the face of waning immunity or novel variants, this tool can be used to support existing processes and qualitative assessments to prioritise countries’ populations for COVID-19 vaccines more consistently, fairly and transparently.

## Methodology

### Study design

The study was conducted in two stages between March and November 2021 and involved public health practitioners and researchers from several countries. The first stage involved a Delphi survey (9) to reach consensus on the most important country-level factors in assessing relative needs for COVID-19 vaccines. The second stage involved a discrete choice experiment (DCE) (10) to determine weights for the factors, reflecting their relative importance. A DCE, also known as conjoint analysis (11), is a survey-based methodology widely used in the social sciences to elicit and understand people’s preferences (informed by their knowledge and expertise). These results were used to create a scoring tool for prioritising countries for access to vaccines.

### Participant selection

Public health experts working in different organisations, countries and specialist areas were identified through the professional networks of three of the authors (VJ, RA, PL). The initial group of participants was chosen *a priori* to ensure they were representative of a variety of institutions and nationalities (Table 1), with the objective of including as wide a range of perspectives as possible. Given the complexity of the study question, experts were chosen purposively, to ensure they had the relevant experience and professional background to contribute. Participants could drop out of the study at any stage, with any earlier contributions retained for analysis. Additional participants were identified through snowball sampling; the first survey concluded with a request to nominate up to five suitable colleagues to participate in the study.

**Table 1.** Included participants.

| Domain | Category | Number of participants (n=28) |
| --- | --- | --- |
| Country/region of focus | Global | 9 |
|  | United Kingdom | 3 |
|  | Japan | 2 |
|  | Kenya | 2 |
|  | Norway | 2 |
|  | South Africa | 2 |
|  | Brazil | 1 |
|  | Chile | 1 |
|  | Mexico | 1 |
|  | Nigeria | 1 |
|  | Peru | 1 |
|  | South Korea | 1 |
|  | Thailand | 1 |
|  | USA | 1 |
| Gender | Male | 17 |
|  | Female | 11 |
| Highest degree (reported) | PhD/DrPH | 15 |
|  | MPH/MSc | 10 |
|  | MBBS/MD/equivalent | 2 |
|  | Other | 1 |
| Job title | Director | 7 |
|  | Professor | 7 |
|  | Consultant/specialist | 4 |
|  | President/principal | 4 |
|  | Senior researcher | 3 |
|  | Assistant Director | 1 |
|  | Senior Advisor | 1 |
|  | Technical lead | 1 |
| Institution | National Public Health Institute | 9 |
|  | University | 8 |
|  | Philanthropic/foundation | 3 |
|  | Department of Health/Ministry | 2 |
|  | Healthcare provider | 2 |
|  | Multilateral organisation | 1 |
|  | Independent charity | 1 |

### Delphi survey

The Delphi survey consisted of two rounds. In the first round, participants were asked to list up to a maximum of ten factors using free text, that they deemed to be most important when assessing a country’s need for COVID-19 vaccines (Supplementary File). Participants were asked to be as specific as possible but only include factors that could be realistically and reliably measured across all countries. Information was also collected about participants’ demographic and professional characteristics, including gender, highest degree, job title, institution and the country in which their work was predominantly based, with a ‘global’ option for people working across multiple countries.

Results from the Delphi’s first round were compiled by the lead researcher (VJ), with all factors reported by at least two participants presented back to the group in the second round. From this combined list, each participant was asked to, in effect, vote on up to eight factors that they considered most important for assessing vaccine needs across countries. All factors chosen by six or more participants were included in the final set of factors for use in the next stage of the research. This quorum of six or more participants was chosen with the objective of ensuring the final set of factors had the endorsement of a significant proportion of all participants (at least 21%, as confirmed in the results).

### DCE survey

To determine weights representing the relative importance of each of the factors included in the final set of factors identified in the Delphi survey, the same group of participants were invited to complete a DCE. The DCE was based on the PAPRIKA method (12) – an acronym for Potentially All Pairwise RanKings of all possible Alternatives – as implemented by 1000minds software (14)(www.1000minds.com). This method and software have been used in a wide range of health applications, including prioritising COVID-19 patients for ICU (13) and hospitalisation (14) and prioritising antibiotic-resistant diseases for research into new treatments (15).

In the context of the present study, the PAPRIKA method involves participants being asked a series of pairwise-ranking questions based on choosing which of two hypothetical countries had the greater need for COVID-19 vaccines (see Supplementary File for examples). The two countries in each question are defined in terms of two factors at a time and involve a trade-off (13) between them (with the other factors assumed the same). A supporting information statement with definitions of the factors and their levels accompanied the DCE. Additional details of the DCE methodology are provided in the Supplementary File.

### Ethics

The experts were initially invited to participate in the study via an e-mail with an information sheet explaining the study background, methodology, risks, benefits and to confirm their willingness to participate. Participants’ data were only accessible by the research team. The study was approved by the UCL Research Ethics Committee (17229/002).

## Results

Initially, 45 experts were invited to participate in the study, and 21 (46.7%) agreed. Snowball sampling delivered another seven participants, resulting in 28 participants in total. They worked ‘globally’ (n=9, 32%.1%) or across 13 different countries (Table 1), with the most frequently reported countries being the United Kingdom (n=3), Japan (n=2), Kenya (n=2), Norway (n=2) and South Africa (n=2). The most common job titles reported were director and professor, with most based in national public health institutes (n=9) and universities (n=8), and most participants’ highest degree was a PhD/DrPH (n=15) or MPH/MSc (n=10).

The first round of the Delphi survey (mid-March to mid-May 2021) resulted in 94 free-text responses identifying factors important in assessing national needs for COVID-19 vaccines, across 28 participants. After aggregating these, 37 factors were identified as having been reported by at least two participants, for inclusion in the Delphi’s second round. They are reported in Table 2, categorised (by the authors) into domains.

**Table 2.**
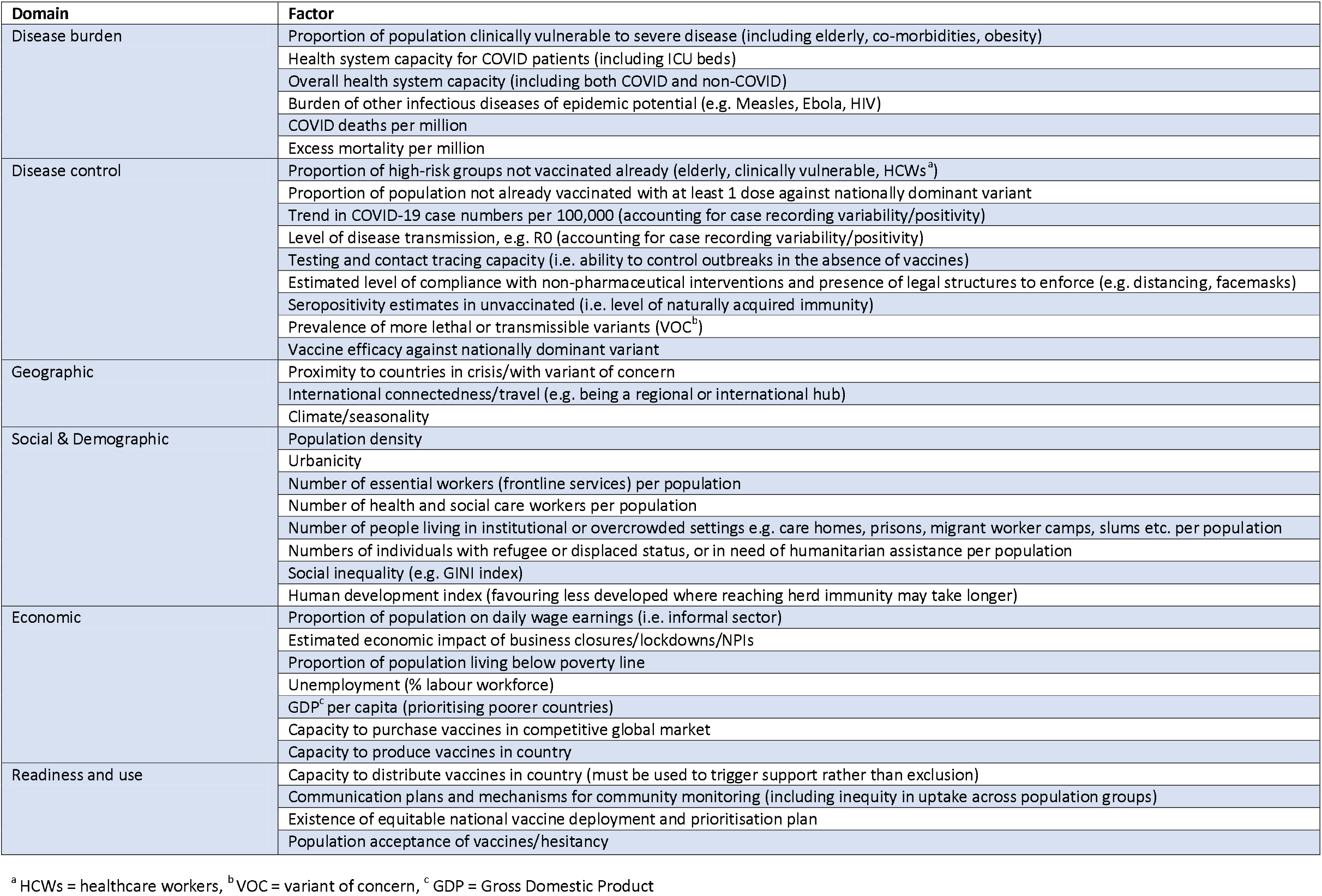
Important factors in assessing national needs for COVID-19 vaccines, Delphi round one results.

Twenty-five of the original 28 experts participated in the second round of the Delphi survey (mid-June to mid-August 2021), which involved each participant voting on up to eight factors from the original 37 factors that they considered most important for assessing vaccine needs across countries. Nine factors received six or more votes, leading to their inclusion in the DCE survey. They and their levels are presented in Table 3.

**Table 3.** Most important factors in assessing national COVID-19 vaccine needs.

| Factor | Overall weight | Levels | Weight by level |
| --- | --- | --- | --- |
| Proportion of overall population not fully vaccinated | 19.5 | Low (10%) | 0 |
|  |  | Moderate (40%) | 12.7 |
|  |  | High (70%) | 19.5 |
| Proportion of high-risk population not fully vaccinated | 16.1 | Low (10%) | 0 |
|  |  | Moderate (40%) | 8.9 |
|  |  | High (70%) | 16.1 |
| Health system capacity | 14.2 | Additional capacity available | 0 |
|  |  | At full capacity | 7.5 |
|  |  | Overwhelmed | 14.2 |
| Capacity to purchase vaccines | 11.9 | High | 0 |
|  |  | Low | 11.9 |
| Proportion of population clinically vulnerable | 11.3 | Low (5%) | 0 |
|  |  | Moderate (10%) | 4.2 |
|  |  | High (25%) | 11.3 |
| Economic impact of lockdowns | 9.4 | Mild | 0 |
|  |  | Moderate | 4.7 |
|  |  | Severe | 9.4 |
| Variant of concern circulating | 7.0 | No | 0 |
|  |  | Yes | 7.0 |
| COVID-19 deaths per million (cumulative) | 6.3 | Low (1000) | 0 |
|  |  | High (5000) | 6.3 |
| National vaccine deployment and prioritisation plan | 4.3 | Does not exist | 0 |
|  |  | Exists | 4.3 |

Of the 25 participants who were invited to do the DCE survey (mid-September to mid-November 2021), 17 at least started it (68%) and 15 completed it (60%), with 36 pairwise-ranking questions answered on average, and most people (12 out of 15) taking less than 24 minutes in total. Only the 15 who completed the survey were included in the final analysis.

The most frequently reported setting of focus for these 15 participants was ‘global’ (n=5), followed by the United Kingdom (n=3). The most common job titles reported were professor (5), consultant/specialist (n=4) and director (3), with most based in universities (n=8), followed by national public health institutes (n=3), and most participants’ highest degree was a PhD/DrPH (n=8) or MPH/MSc (n=7).

The heterogeneity of the 15 participants’ preferences is illustrated by the radar chart in Figure 1, representing each participant’s individual weights, and their means. The mean weights are reported in detail, including for their levels in Table 3. These factors and levels and their weights constitute the prioritisation scoring tool.

**Figure 1.**
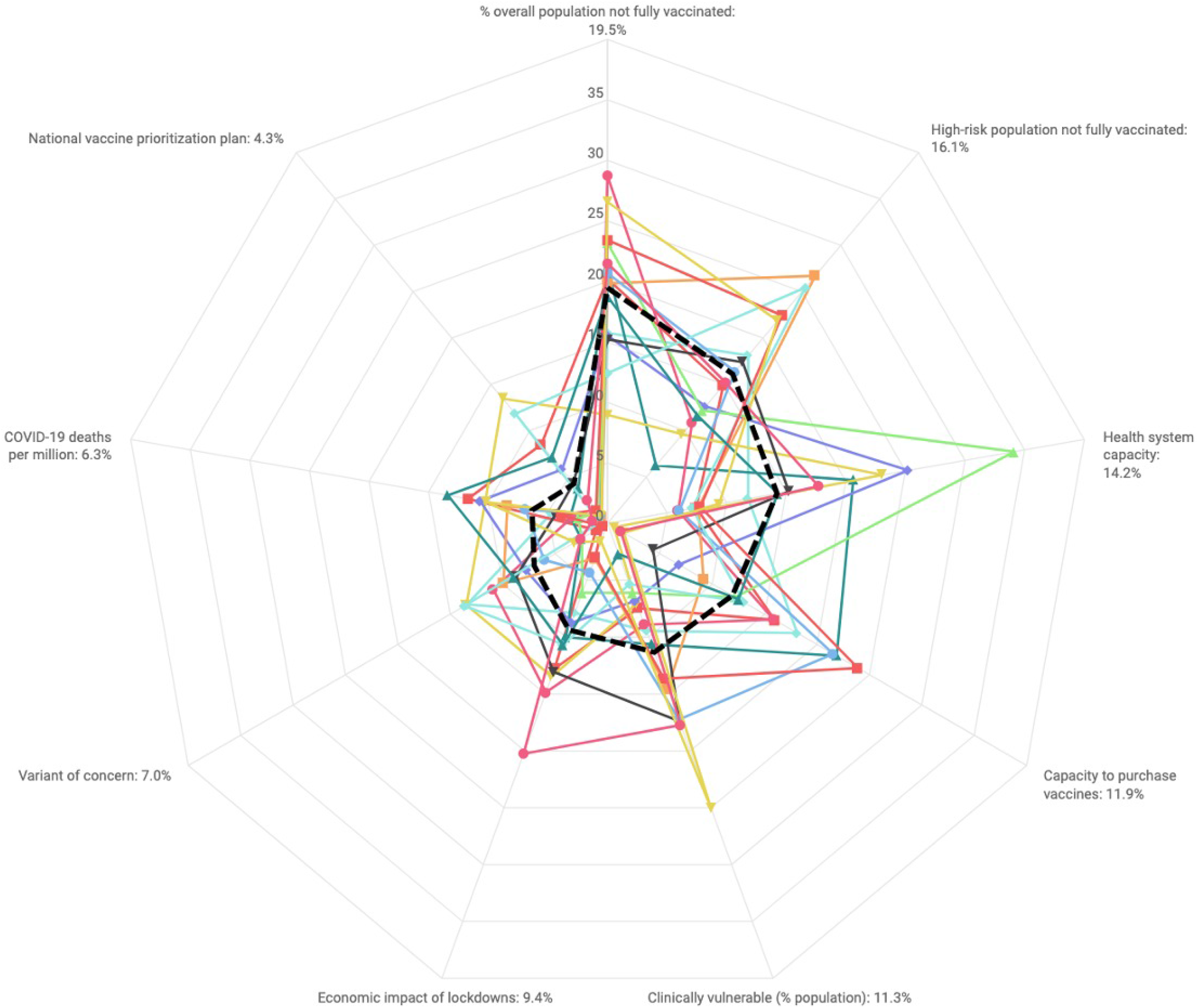
Radar Chart of Participant Responses in DCE (Black Dashed Line = Mean Weight)

As can be seen in Table 3, the most important factor for assessing vaccine needs across countries was ‘proportion of overall population not fully vaccinated’ with a mean weight of 19.5, followed by ‘proportion of high-risk population not fully vaccinated’ (16.1), ‘health system capacity’ (14.2), and so on, down to – the least important factors – ‘COVID-19 deaths per million (cumulative)’ (6.3) and ‘national vaccine deployment and prioritisation plan’ (4.3). These criteria and levels and their weights constitute the prioritisation scoring tool.

Fourteen (93%) of the 15 DCE participants agreed that the ranking of factors produced by their DCE seemed roughly correct to them; 12 (80%) said the survey design was not difficult to understand; and 10 (66.7%) agreed that the survey included the most important factors relating to national needs for COVID-19, with 3 (20%) stating that they were not sure.

## Discussion

Several distinct factors were considered important in assessing national needs for COVID-19 vaccines across countries, including factors related to disease burden, disease control, health system capacity, geographic, social, and demographic, economic and readiness.

On average, participants considered the proportion of the overall population and of the high-risk population not fully vaccinated to be the most important factors. These two factors were four to five times more important than the least important factor included in the DCE survey, the existence of a national vaccine deployment and prioritisation plan. Factors not routinely used in real-world global vaccine allocation (3,6), but deemed important in this study, included the proportion of the high-risk population not vaccinated, the economic impact of lockdowns, variants of concern, COVID-19 deaths, and the existence of a national vaccine deployment and prioritisation plan.

### Relevance to the ongoing COVID-19 pandemic

Existing vaccines are less effective in preventing symptomatic disease and transmission with the Omicron variant compared to previous variants, with booster doses providing additional protection against severe illness (18). The global shift towards booster doses in response to Omicron is exacerbating global vaccine inequity (19). Boosters can only be given after a primary course of vaccination, with several countries recently shortening the interval at which individuals are given a booster (20).

Though the World Health Organization (WHO) has been advocating for countries to vaccinate 70% of their populations (with a primary course) by the middle of 2022, at the current pace, 109 countries will likely miss this target (21). Our findings will support the fair allocation of primary courses of vaccines across countries, which in turn may limit the inequity caused by the disproportional procurement of booster doses by wealthy countries (7). Given the possibility for regular COVID-19 vaccine boosters, and the threat of novel variants requiring a further tweaking of vaccines, this scoring tool represents an important step towards equity in long-term pandemic response.

### Assessing vaccine needs across countries

Needs assessment for COVID-19 vaccines at the country-level is complex but the COVAX Facility aims to deliver vaccines to cover 20% of all populations, including the elderly, those with co-morbidities and healthcare workers. After this, countries receive doses based on need, based on: the effective reproduction number (R number) and its trend, hemisphere location, universal health coverage (UHC) service coverage index, health system saturation, and the size of groups at a high-risk of severe disease or death, supported by a qualitative assessment (6). In addition, up to 5% of vaccine doses are reserved as part of a humanitarian buffer for populations such as refugees or asylum seekers. All these factors were identified in our Delphi survey, but not all were among the most frequently reported factors.

The Fair Priority Model is an alternative tool (5), which uses reproduction numbers, years of life lost and national economic indicators to help consider vaccine needs, with needs assessment changing over time with sequential vaccine programme objectives. A third tool, proposed by researchers at Vanderbilt University (2), suggests allocating vaccines to countries based on their ability to distribute vaccines and capacities to provide care, which we identified as one of the most important factors relating to vaccine needs.

Despite a range of proposed tools to allocate scarce vaccines across populations, all involve the use of a relatively narrow set of metrics and fail to assess or account for the relative importance of the various indicators included. In order of decreasing importance, the factors not considered by COVAX (6) which were deemed among the most important by participants in our study included the proportion of the high-risk population not vaccinated, the economic impact of lockdowns, variants of concern, COVID-19 deaths, and the existence of a national vaccine deployment and prioritisation plan.

Our previously proposed framework (COVID-NEEDS) (8) included six of the nine most important factors identified in this study. The added value of the proposed scoring tool will likely be in expanding discussions to better consider horizontal equity, where country needs are deemed to be similar according to existing risk assessments based on a less comprehensive set of indicators. Many rich countries have used vaccines as a form of foreign aid (22) tied to diplomatic or economic objectives. Our findings, by making national vaccine needs assessment more transparent, explicit and objective, may help to increase the role of equity and minimise the role of politics in such decisions.

### Operationalising the prioritisation scoring system

To consolidate and regularly update data on the identified factors in a single tool for all countries will be challenging. Nevertheless, this is possible given the wealth of COVID-19 data available in the public domain and the ability of international institutions such as the WHO to access further real-time information at the country-level. The assessment of some factors (e.g. size of the clinically vulnerable population) will be based upon estimates, which though available across countries (23), may vary in quality. An assessment of capacity to purchase vaccines can broadly be considered by country income-group status, with low-income countries having a very limited capacity to compete with wealthier ones with respect to ability-to-pay for vaccines from manufacturers (24).

The available quantitative data (including an understanding of variation in quality) must be considered alongside qualitative information from stakeholders within countries and familiar with real-time on-the-ground realities. This may be particularly important to support some quantitative metrics such as on COVID-19 variants, due to the risk of under-prioritising countries with limited genomic surveillance capacities (25). Current COVAX plans propose using both qualitative and quantitative data for the same purpose (6), meaning the current scoring tool will not require any additional ancillary inputs above those within existing WHO processes.

For the domestic allocation of COVID-19 vaccines, most countries have opted for relatively simple methods (26,27). This has increased the speed at which populations have been immunised. For international allocation, speed and logistics have been similarly important as reflected in COVAX plans. Further research is required to understand how the use of this scoring tool may affect the efficiency of real-world vaccine allocation. As with many health interventions, there may be a trade-off between equity and efficiency (28). But because most of the factors in our scoring tool are part of existing international risk assessments, it is unlikely to have a substantial impact on the efficiency of international vaccine allocation.

### Limitations

Although we were able to include a wide range of experts in this study, our findings may not necessarily be representative of all experts in the field. Having said this, the tool improves on existing priority-setting mechanisms. Factors were only included if reported by multiple experts working across several countries and institutions, and determining their weights was performed using a choice-based exercise (the DCE) instead of more traditional questionnaire or ranking methods.

In addition, the study was limited by a response rate of approximately 50% of the experts we invited. Although all participants initially agreed to participate, most of them were actively contributing to the pandemic response, leading to significant time pressures, meaning that many were unable to complete the DCE. The mean weights for some factors may have been different with additional participants, although large changes would be required for the ranking of the factors to change substantially.

The study also started before booster doses were widely used, meaning that the factors and weights identified may differ for the exclusive allocation of booster doses, given differences in epidemiological utility for disease control and severity compared to primary course vaccination (18,29,30).

Finally, we were limited in the amount of contextual detail we were able to provide in the DCE survey. There may have been further factors, beyond the two outlined for each country, that influenced decisions about which country was in greater need for COVID-19 vaccines. But by assuming everything else remained constant, we were able to isolate the quantitative importance of specific factors, which can further support more qualitative country-specific information.

### Implications for research

To further develop the evidence-base, future studies must aim to compare needs for vaccines as assessed by experts to those of decision-makers and members of the public. Given the vast range of impacts of the COVID-19 pandemic, it is difficult for any single group of individuals (including experts) to provide a comprehensive understanding of the range of factors involved in assessing needs for vaccines. Although several technical issues may be better understood and analysed by experts, the value judgements of all stakeholders involved (including the public) must be considered, to develop robust, inclusive and sustainable priority-setting processes (31,32).

Disease X (as it has been coined) represents the knowledge that a serious international pandemic could be caused by a pathogen currently unknown to cause human disease (33). Although our findings cannot be directly extrapolated to future health emergencies of different diseases, they highlight the complexity of international vaccine needs assessment. Several of the factors identified here may prove to be important considerations for future crises, and by making them explicit, our study will aid international discussions in the early phases of response to the next public health crisis.

## Conclusions

Assessing needs for COVID-19 vaccines is complex, given the extensive but variable impacts of epidemics across populations and the diversity present in social value judgements. Several factors exist, extending beyond traditional metrics, which may lead to particular countries having a greater need for vaccines compared to others. On average, the proportion of the overall population and of the high-risk population not fully vaccinated, were the most highly valued factors related to vaccine needs. Several other factors found to be important, such as the economic impact of lockdowns, are not routinely considered in global vaccine allocation mechanisms. This scoring tool will aid qualitative assessments to further the role of equity in global vaccine allocation.

## Data Availability

Survey data collected and used in the analysis from participants are confidential and not routinely available. Anonymized data may be made available to other researchers upon reasonable request and author review.

## Authors and Contributors

All authors contributed to all stages of this study, including inception, design, literature search, data analysis, data interpretation, illustrations, write-up and discussion, editing and revisions.

Substantial contributions to the conception or design of the work; or the acquisition, analysis, or interpretation of data for the work: VJ, PL.

Drafting the work or revising it critically for important intellectual content: VJ, PH, RA, PL. Final approval of the version to be published: VJ, PH, RA, PL.

Agreement to be accountable for all aspects of the work in ensuring that questions related to the accuracy or integrity of any part of the work are appropriately investigated and resolved: VJ, PH, RA, PL.

## Acknowledgements

We would like to thank all involved individuals for their valuable time, expertise and participation in this project, without whom this study could not have been completed.

## Tables and Figures Legend

Table 1. Included participants

Table 2. Factors associated with national needs for COVID-19 vaccines, Delphi round one results

Table 3. Most important factors related to national COVID-19 vaccine needs

