## Supplementary material for "Which countries need COVID-19 vaccines the most? Development of a prioritisation tool": Supplmentary File

**Supplementary File S1**

**Contents**

Delphi Survey Questions........……………………………………………………................…………………………...............................………………………………2

Examples of the DCE’s Trade-off Questions…………………….……………..…….………………………………...........………………............….…………………3

Preference Elicitation Validation Questions....................………………..……………………….……...........………………………………….…………….………5

Preference Elicitation Supporting Information Statement.……………..……………………….……...........………………………………….…………….………6

DCE: Additional Details…………..……………………….……...........………………………………….…………….……………..……………………….……...........………7

**Delphi Survey Questions**

First Round

Please list up to a maximum of ten factors that you would deem most important in an assessment of national needs for COVID-19 vaccines. This may reflect the impact of COVID-19 itself as well as associated non-pharmaceutical interventions. Please be as specific as possible but include only factors that can be realistically and reliably measured at the country-level. You do not need to describe exactly how you would measure your proposed factors in this round of the survey. Proposed factors relating to vaccine need should exclude existing political and diplomatic considerations, instead aiming to reflect an ideal, fair and scientific assessment of vaccine needs. Your ideas will help to build on currently proposed indicators of the COVAX Facility Vaccine Allocation Mechanism: <https://www.who.int/publications/m/item/fair-allocation-mechanism-for-covid-19-vaccines-through-the-covax-facility>

Second Round

### All the factors below [factors listed in answer options] were identified as important national-level factors in assessing needs for COVID-19 vaccines, by at least two survey participants. To establish consensus on the most important of these, please choose up to 8 of the following factors that you believe are the most useful in forming a well-rounded assessment of needs for COVID-19 vaccines across countries. These should tell us whether one country needs vaccines more urgently than, or ahead of, another. Please prioritise factors that you believe are not just most important but also feasible to measure and routinely available at the national level across all countries.

**Preference Elicitation Example Questions**

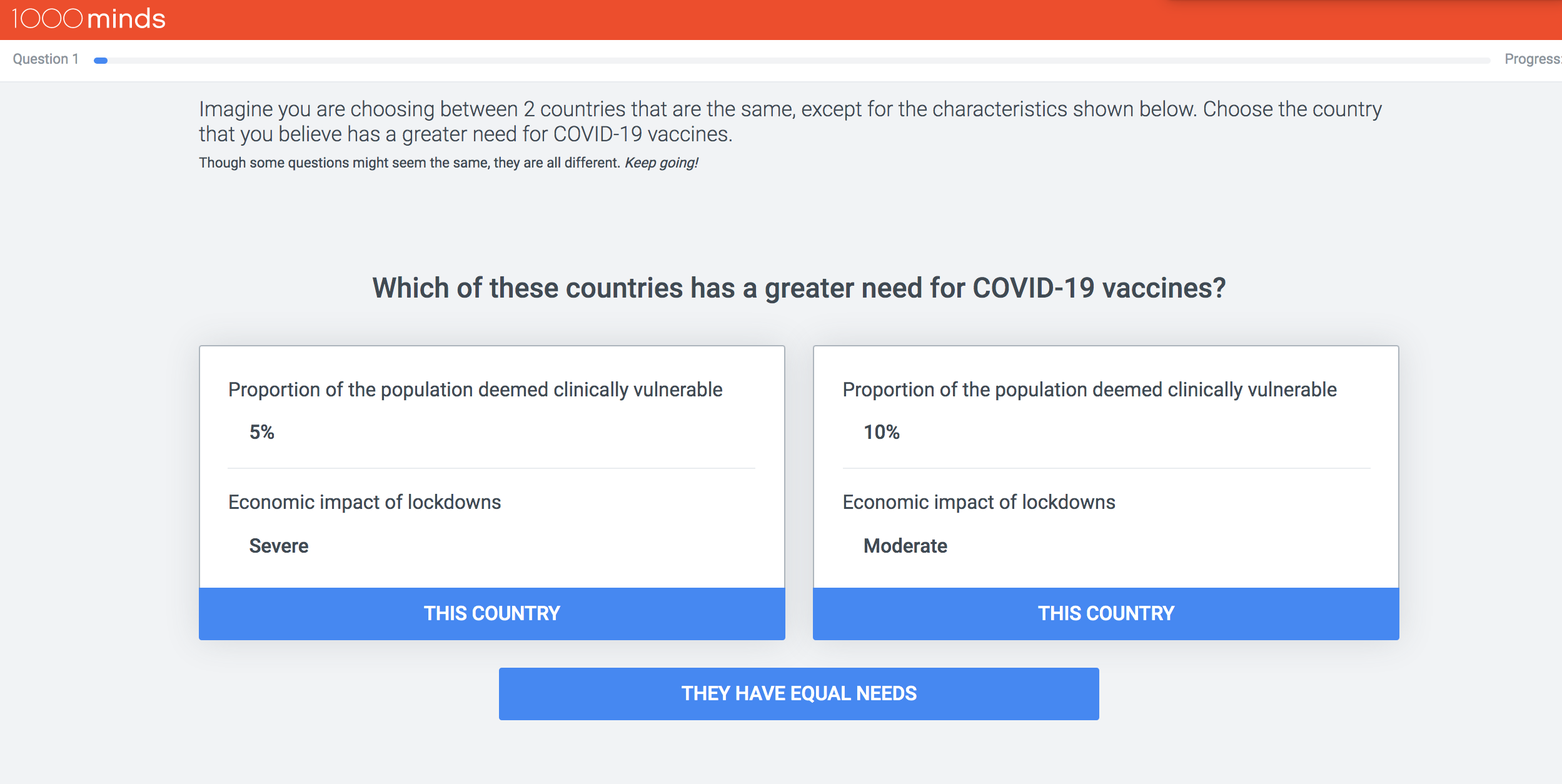

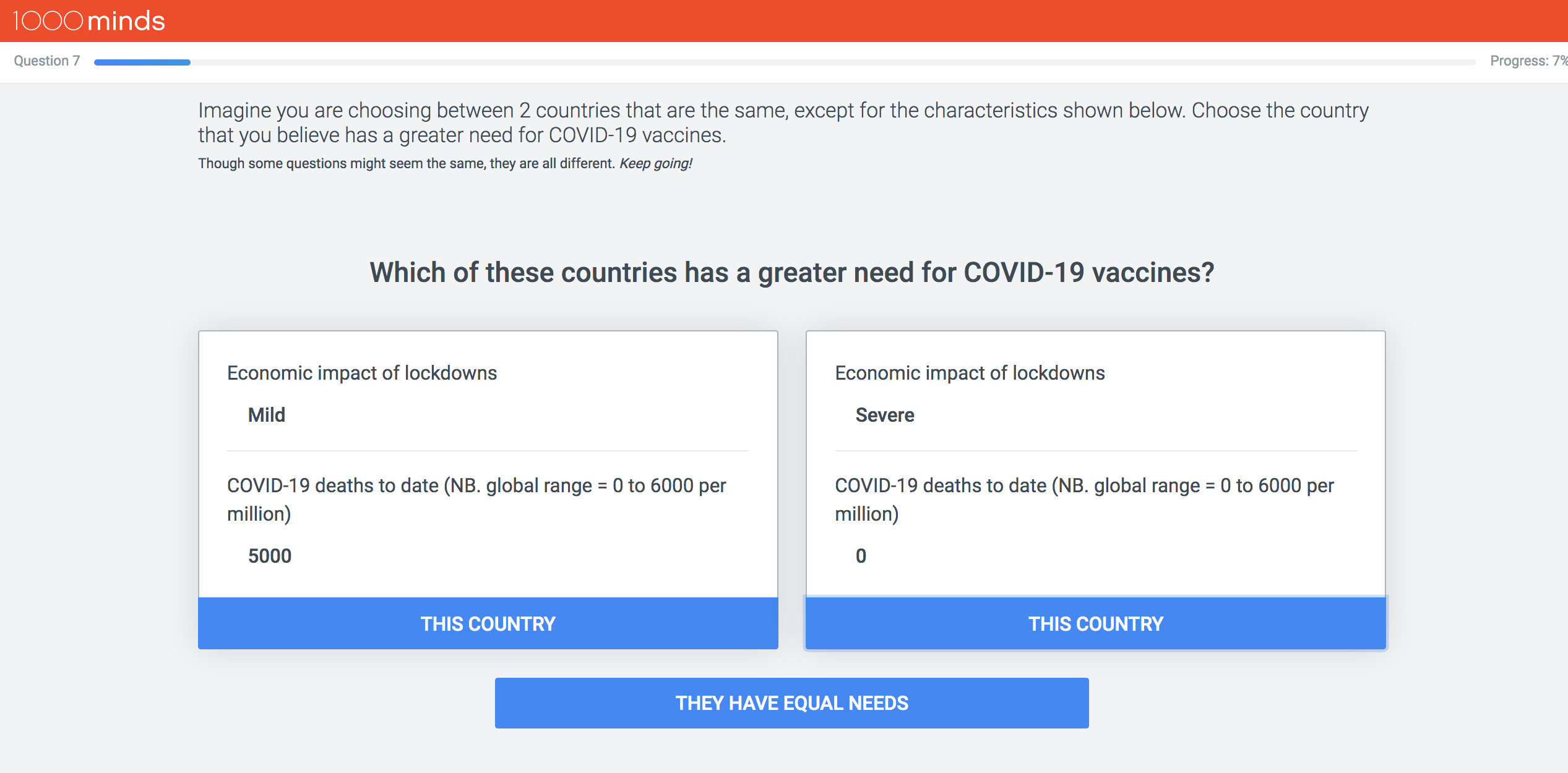

**Preference Elicitation Validation Questions**

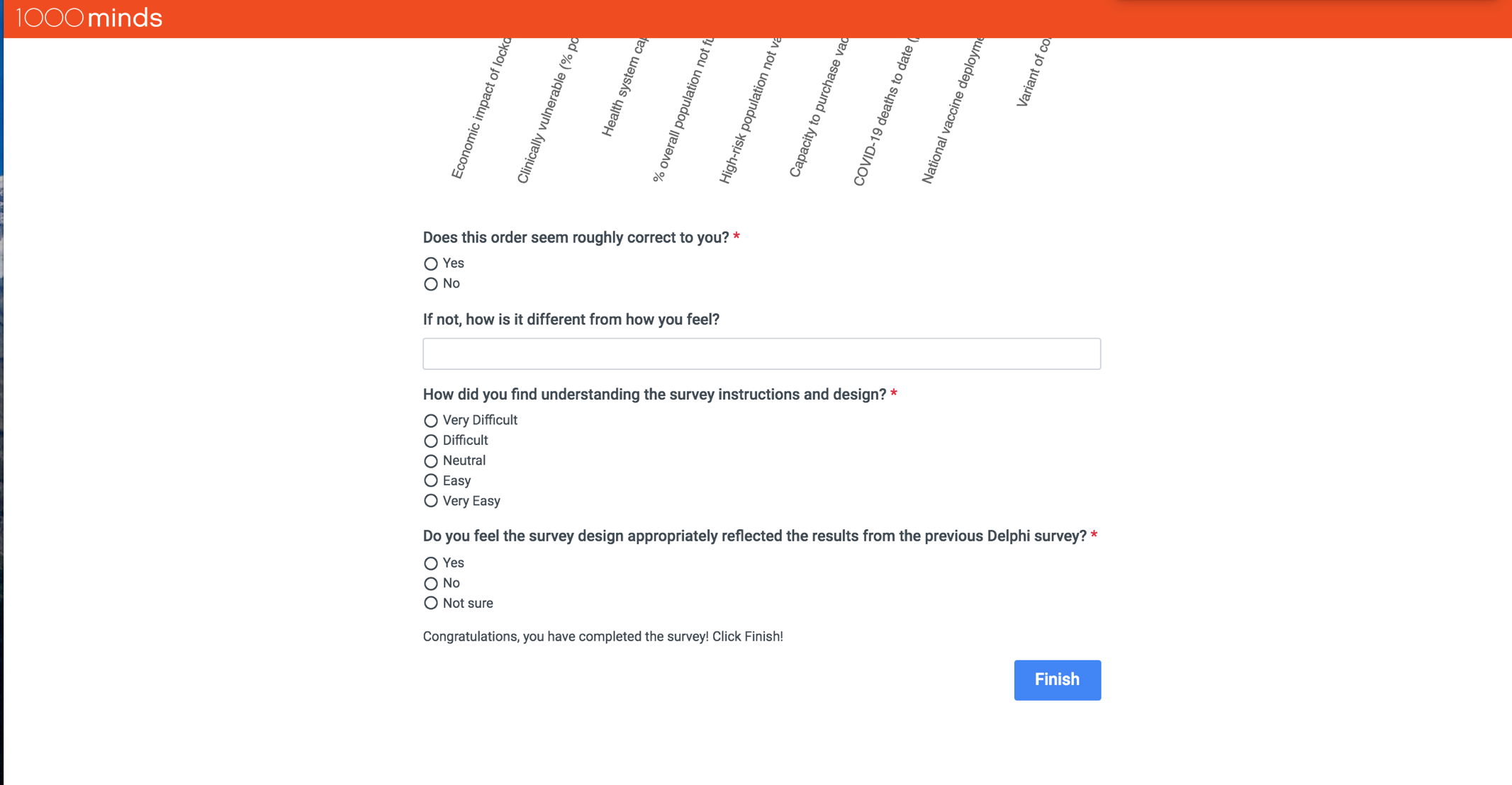

**Preference Elicitation Supporting Information Statement**

Factors predicting needs for COVID-19 vaccines:

Proportion of clinically vulnerable (e.g. elderly, those with co-morbidities) not fully vaccinated

- 10%

- 40%

- 70%

Proportion of population who are clinically vulnerable (e.g. elderly, those with co-morbidities)

- 5%

- 10%

- 25%

Economic impact of lockdowns (i.e. stringent restrictions on mobility, social interaction and access to public spaces)

- Mild

- Moderate

- Severe

Health system capacity

- Additional capacity available

- At full capacity

- Overwhelmed

COVID-19 deaths per million (NB. global range = 0 to 6000), from the start of the pandemic until now

- 1000

- 5000

Variant of concern circulating (i.e. more transmissible and/or lethal, unknown susceptibility to vaccines)

- No

- Yes

Proportion of population not fully vaccinated

- 10%

- 40%

- 70%

Capacity to purchase vaccines (in competitive global markets)

- Low

- High

National vaccine deployment and prioritization plan

- Does not exist

- Exists

**DCE: Additional Details**

For each participant, the pairwise-ranking questions are repeated with different pairs of hypothetical countries (always defined on two factors at a time and involving a trade-off). Each time the participant makes a choice, the method adapts by formulating a new pairwise-ranking question based on all the participant’s previous choices. This adaptivity, which serves to minimise the number of questions each participant needs to answer, is based on the method’s application of the logical property of ‘transitivity’. For example, if hypothetical country *A* is preferred to country *B* and *B* is preferred to country *C*, then, logically – by transitivity – *A* is also preferred to *C*, and so a question about this third pair is not asked.

From the participant’s answers to the pairwise-ranking questions, the software uses quantitative methods to derive weights (sometimes called ‘utilities’ in the DCE literature) for each of the factors, representing their relative importance. As well as individual weights for each participant, mean weights are reported for the group. These mean weights are used as scores (between 0 and 100) representing the relative importance of each factor (and level).

Upon the DCE’s completion by each participant, as a face validity check, they were shown their weights and asked if the ranking of the factors seemed “roughly correct to you”, and if not “how is it different from how you feel?”. Finally, a question was asked about how easy or difficult they found understanding the survey instructions and design, and whether or not they felt the DCE included the most important factors relating to national needs for COVID-19 vaccines.

The survey was pilot tested with seven public health experts known to the researchers but who did not participate in the survey. Based on that feedback the survey was refined, and then a link to the survey was emailed to participants.
